# Proposed standards for prosthetic foot reuse and considerations for donation of used prosthetic feet to low-and middle-income countries

**DOI:** 10.1101/2025.08.01.25332618

**Authors:** Michael A Berthaume, Louise Ackers, Laurence Kenney, Vikranth Harthikote Nagaraja, Promise Maduako

## Abstract

Prosthetic limb provision in low-and middle-income countries (LMICs) is complex. Prosthetics from high-income countries (HICs) are often replaced not because they are broken, but because of warranties and/or guidelines, meaning they may still be usable. However, as there are no standards/regulatory requirements guaranteeing the quality or safety of second-hand devices, they are often disposed of or donated to LMICs. Without standards to guarantee the quality of donated prosthetics could lead to a violation of the World Health Organization’s principles of good donation. Here, we compare 196 feet delivered prior to the introduction of an internal check for prosthetic foot quality introduced by a UK-based charity (STAND) with 170 feet delivered afterwards. The implementation of the check increased the percentage of usable second-hand prosthetic feet received by donation recipients from 83.16% to 94.12%. Foot brand significantly affected usability, but further data and larger samples are needed to disentangle the effects of prosthetic brand, prosthetic foot model, and donation centre on prosthetic foot usability. We propose a rapid and efficient quality checklist as a set of standards governing used prosthetic foot reuse and donation and discuss future directions for this line of work. The creation of an international set of standards/regulatory requirements governing the use of prosthetic limbs, like the international standards used for prosthetic limb design, would not only enable the safe, useful provision of prosthetics in LMICs, but would also set the groundwork for understanding how a repair, reuse, and recycle model for prosthetics might be implemented in HICs. Globally, this could decrease prosthetic provision time, create a circular economy for prosthetics, and reduce the carbon footprint of prosthetic manufacture and provision.

## INTRODUCTION

Despite being a priority assistive product and a basic human right (Convention on the Rights of Persons with Disabilities; World Health Organization, 2016) and critical for the sustainable development goals (SDG 3: Good Health and Well-Being), poor levels of access to prosthetic limbs are common in low-income and low-and middle-income countries (LICs, LMICs). Systematic issues arise in terms of funds (e.g., to purchase prosthetic devices, or pay for repairs), access to professionals (prosthetists, technicians, etc.) and materials (e.g., polypropylene), and access to prosthetic and orthotic (P&O) centres by the patients, which are exacerbated in rural areas [2]. Various methods of prosthetic provision have arisen to meet these challenges.

### Prosthetic provision in LMICs

Prosthetic provision in LMICs involves inter- and intra-national stakeholders. This is particularly true during times of conflict, when amputees are a visible reminder of the effects of conflict on the person, and the media and public draw a moral imperative, empowering many stakeholders to help [3]. Stakeholders include non-profit organizations (NGOs), charities and faith-based organizations (FBOs), for-profit and hybrid for-profit/NGOs, prosthetic training centres, and hospitals/P&O centres themselves [2]. This creates a mostly “top-down” approach from high-income countries (HICs) to LMICs [4], but bottom-up approaches, such as Community-Based Rehabilitation (CBR), exist. CBR utilizes local social and community infrastructure and provides some benefits (Cummings, 1996), particularly in creating amputee support networks [5].

Provision of prosthetic devices includes prosthetic hardware, (locally) manufactured (bespoke) sockets, and the surrounding systems and services, such as local skillsets and supply chains [6]. When providing prosthetics, cultural, economic, social, phycological, religious, and climatic factors, as well as the integration into local systems, should impact the choice of technology/services provided [4]. Poonekar suggested LMIC prosthetics should be (i) low-cost; (ii) locally available; (iii) capable of manual fabrication; (iv) considerate of local climate and working conditions; (v) durable; (vi) simple to repair; (vii) simple to process using local production capability; (viii) reproducible by local personnel; (ix) technically functional; (x) biomechanically appropriate; (xi) as lightweight as possible; (xii) adequately cosmetic; and (xiii) psychosocially acceptable [7].

Cultural and sociocultural norms are often overlooked but can be particularly important. For example, karma may lead a people to believe amputees lost limbs because of cosmic balance and can lead to an indifference in amputee aid, whereas the “alms for the poor” mentality can lead to larger community engagement and aid [3]. Heim (1979) discusses case studies where a lower-limb prosthetic user refused a ‘peg-leg’ because other users did not have peg-legs and they wanted to be treated as an equal. Similarly, when patellar tendon bearing (PTB) prosthetics were being given out in a tribe, one man – who was not given a PTB socket – wanted one to be like the others [8]. Conversely, the introduction of new technologies can be difficult to accept due to anxiety about the technology’s performance or embarrassment about using the technology for the first time, particularly among first-time prosthetic users [9]. Acceptance can increase once users see proof new technologies work well with others [8]. If assistive devices fail to meet cultural needs, they can be abandoned [10–12], although the extent of device abandonment in LMIC settings is an under-researched area. “… unsuitability of assistive devices for the topographical and cultural (social and attitudinal) contexts in many developing countries, which are then strongly linked to poor patient outcomes, including dissatisfaction and limited participation in meaningful activities [13].” Within these contexts, it is important to remember culture is fluid and ever changing. The interdisciplinary approach combining anthropological (e.g., cultural) and engineering concepts can lead to the improved design and provision of prosthetics [14–16].

Despite the best of intentions, prosthetic provision does not always go well [17], and the provision of inappropriate technologies or services can cause harm [5]. Ignorance of social/environmental issues, as well as grouping of all LICs/LMICs, developing countries, and/or low-resource settings into the same categories, is problematic. “The commonalities of poverty, lack of resources, demographics and the prevalence of diseases, which have been eradicated in the rest of the world, can lead to sweeping generalisations regarding the needs of LICs. Unique aspects of each country’s history, politics, culture and social structure must be considered when planning any prosthetic and orthotic service [18].” Prosthetic providers may wish to provide a large range of products to a single end-user group to ensure all end-user needs are met. However, this approach may not be practical, as it requires sustained access to a broad range of supplies and skillsets [8].

There are, however, some common issues with prosthetic provision across all LMICs. Charity-based and globalisation models are the most prominent models of assistive technology provision in LMICs [6,19]. Bulk manufacturing can lead to a decrease in assistive technology cost, but inadequate buying power by providers in LMICs can prevent them from realizing these benefits [20]. Bulk production can also lead to a limited range of prosthetic technologies available [20] – rather than choosing the best prosthetic technology/service for a population, prosthetic technologies/services are presented based on availability to/by the organisation [3]. Even when choice is available, once in country, products are often provided on a first-come first-serve basis as materials are received instead of the most appropriate technologies being given to each person [5]. As globalization, which tends to focus on money spent or number of prosthetics given and not how the user’s life has improved [5,21], can disrupt local supply chains and squash local innovation [6,19], it can be difficult to develop and/or provide devices and services which fulfil end-user needs.

### The donation model of prosthetic provision

The donation model for prosthetic provision – which overlaps with the reuse and repair models [6,22] – is embedded within FBOs, NGOs, and for-profit/hybrid organizations [2], where it is the responsibility of the donor to ensure device quality and safety are met. It operates in a complex ecosystem which involves a diverse set of stakeholders, who often navigate numerous barriers affecting quality and appropriateness of donations, donation sustainability, and donation optimization [23].

If new or second-hand products meet standards and regulatory requirements, they can be a major supply source for some countries, providing advanced, otherwise unavailable technologies for cheaper-than-market prices [6,20]. While donated medical devices typically meet safety, quality, and performance requirements, Nasir et al. (2023) found regulatory and guidelines/processes for donated medical devices in African countries to be inadequate. This is partially due to oversight and ill-defined/poorly understood medical device regulations. Similar issues have been noted elsewhere (e.g., [24–26]).

Standards for reuse, repair, and donation of devices can exist at the organization, country, and international level. For example, an organization may only donate devices that reach certain internal quality standards, while governments and international organizations, like the World Health Organization (WHO), may set their own standards or regulatory requirements. At the Regulatory Framework for Assistive Devices in Uganda, organized by the Ministry of Health, there was a plea to ‘tell people to stop dumping their junk on us’, highlighting the need for quality standards which are set and adhered to by international organizations. Several checklists, guidance, and frameworks have been established to standardize donation of medical devices and equipment, highlighting the need to consider the donation process holistically [23,27].

When donating, it is good practice to adhere to WHO principles of good donation, although it is important to recognize they are not international regulations, but guidelines meant to help national and institutional organizations dealing with health care equipment donations create their own standards/regulations [28]. These include 1) ensuring health care equipment benefits the recipient to the maximum possible extent, 2) providing donations that are desired and fit local standards, 3) ensuring there are no double standards in quality, and 4) effective communication between donor and recipient while ensuring agreed upon donation plans are adhered to. Donators should work with local stakeholders and coordinate with national service systems, ensuring consistency of service, follow-up, data control, choice of technology, trained personnel, and sustainability [27,29,30]. The provision of product specifications and use guidelines in local languages aids with technological sustainability is the goal [29,31]. Only working/needed equipment should be donated, and recipients – be that countries, organizations, or device users – should be allowed to turn down donations. Time and budget should be allocated to follow-up on donations, e.g., for maintenance and repairs [31], and donators should stay involved for years for the establishment of prosthetic services, to ensure longevity and sustainability [8].

Despite distinct advantages with the donation model, issues exist with sustainability, quality, compatibility, infrastructure, and training [32], and the donation of medical equipment can unfortunately have adverse effects [31,33]. In the surgical/anaesthesiology sectors, “Lack of planning and collaboration can mean that equipment donated with ‘good intentions’ to help address these shortages is inappropriate, ineffective or dangerous [31].” Even when donations are high-quality and appropriate, differences in training and reliable infrastructure availability can mean technologies that work well in HICs work poorly/are unsustainable in LMICs [31]. Within assistive technologies, donated devices can fail users in the short-term by having inconsistent supplies, being of low quality, and/or being inappropriate for the context of use, e.g., not working well in the rugged environments of the target countries [2,6,20,29]. For example, “Donations from [overseas] of used hearing aids, they used us as a dumping ground, they were not functioning well [29].” In the long-term, failure can negatively affect quality of life in LMICs [13], and shifts responsibility for the disposal of medical devices to already overstretched local services. Medical device companies are not obliged to translate their documentation into languages of donation-recipient countries, and the lack of doing so is bad practice [29,31] and inevitably causes challenges, particularly with the more complex devices [34].

Donated items come with reduced lifetime and performance and increased servicing and maintenance challenges [11]. Products can be difficult to repair as it may not be possible to obtain replacement parts – putting reliability of future supply on the donors [19] and sometimes meaning recipient countries re-use single-use items – and/or personnel may not have proper training [31]. “Even when projects utilise donated western devices, their complexity predisposes then to require regular repair and replacement. It is difficult to refuse free devices even when they are not fit for purpose [18].” While donated products/services provide immediate relief (assuming they are not abandoned by the user [5]), they can lead to dependency on external sources, preventing the establishment of local supply chains and hindering the development of local manufacturing and maintenance capabilities [35]. Ultimately, good intentions are not enough; donors should consider human resources, environment, material resources, maintenance resources, and education resources when donating prosthetics [31].

### Donation of prosthetic lower limbs and feet

There are no international standards and regulatory requirements governing the use of used prosthetic limbs or their componentry, and there is limited work on P&O donation in LMICs [36], meaning there is no way to guarantee their quality or safety when reused. Consequently, many HICs/companies classify prosthetics as single-patient multi-use medical devices which can be repaired and given back to the same patient when in warranty, and the inclusion of out-of-warranty used prosthetic parts in new prosthetics is not allowed. However, as within-warranty prosthetics are regularly repaired and provided back to patients, it is clear prosthetics can be repaired and reused. As many prosthetic components are overdesigned, there is still significant (if, as yet, unquantified) life left in at least some prosthetic components at the time of replacement.

Many organizations collect and donate lower-limb prosthetics to LMICs. For example, Humanity & Inclusion’s Liimba project (launched in 2006) collects and refurbishes used prostheses from Belgium, France, Luxembourg, and Switzerland – where prosthetics are replaced every five years but cannot be reused within country – for donation in countries where Humanity & Inclusion operates (https://www.hi.org/en/liimba--giving-a-second-life-to-prostheses-thanks-to-refurbishment). Similarly, Limbs for Life Foundation collects and refurbishes used prosthetics and prosthetic parts from the United States – which again are usable but cannot be reused within country – and provides prosthetics/componentry to other countries through partnerships, including Human Engineers Inc. (Philippines), Thomas L. and Linda J. McCormack Foundation (Panama), Protesis Imbabura (Ecuador), 2ft Prosthetics (Dominican Republic, Mexico, Philippines, Tonga), and STAND (sub-Saharan African) (https://www.limbsforlife.org). Another US organization, Hope to Walk, provides its own prosthetic legs for LMIC contexts, but also accepts and donates recycled prosthetic legs and parts (“If you would like to donate old prosthetic legs and parts to be used for these patients, please <u>contact us</u>.” https://www.hopetowalk.org).

STAND, formerly Legs4Africa, is a UK-based charity that has collected 14,000+ donated, used prosthetic legs from individuals and P&O centres across US, Canada, UK, and across continental Europe. For feet to be accepted by STAND, they must have no visible cracks, wear, or degradation and be complete with no missing parts. They also must be modular prosthetics, 1) ensuring easy dis- and re-assembly, 2) allowing for multiple brands and types of technology to be combined, and 3) enabling greater flexibility in alignment/adjustment throughout the prosthetic fitting/repair process for the prosthetists in the network. Prosthetics collected in the US, Canada, and UK are shipped to Bristol where they are disassembled into their components (sockets, pylons, feet, and/or knees), inspected, boxed, and relevant parts are shipped to partner clinics in western and eastern Africa. In eastern Africa, components are shipped to Dar es Salaam, Tanzania, and redistributed to rehabilitation centres in Kenya, Tanzania, and Uganda. Prosthetics collected in continental Europe are shipped to Nav Solidaire in Normandy, France; Nav Solidaire is STAND’s collection partner in France and performs similar activities to STAND in Bristol (visual inspections and cleaning of the components). However, their operation involves shipping the components southwest across the Atlantic to The Gambia and Senegal. A carbon impact assessment conducted by an independent assessor estimated STAND saves 5.7+ tonnes of CO2e emissions/year by rescuing compared to manufacturing new parts.

As there are no standards and regulatory requirements for used prosthetic componentry, STAND initially disassembled modular prosthetics and quickly visually inspected components for damage. However, some components – mostly feet and foot shells – would break during or shortly after shipment. To improve quality, prosthetic feet underwent additional scrutiny from January 2023 onwards, including:

1. Checking feet were complete and bumpers were present,
2. Inspecting for surface flaws including cracks, degradation, or discolouration that may affect cosmesis or indicate the onset of foot deterioration,

a. During surface inspection, toes are bent and thumbs were pressed into the foot shell to reveal any hairline cracks that would otherwise be invisible.
b. Toe flexibility is also noted when toes are bent to ensure the toes are not overly flexible.
3. Ensuring non-SACH feet have matching keels and shells (size, model, and brand)

If the feet fail any one of these criteria, the foot is deemed unsuitable for donation and discarded.

As a first step to understand what type of international guidelines, standards, or regulations may be required for the reuse of prosthetics, we examine the efficacy of the organizational standards used by STAND, with the idea that proven efficacy of these standards could lay the groundwork for the development of future international standards. Our underlying scientific hypothesis is that the implementation of these standards will lead to an increase in the proportion of usable prosthetic feet. To include any damage that could occur during shipping, we review prosthetic feet already in Africa.

## MATERIALS AND METHODS

In February 2025, MB and PM travelled to Fort Portal, Uganda and reviewed prosthetic feet donated by STAND. 196 feet were stored at the Ninsiima Centre for the Rehabilitation of People with Physical Disabilities (NCRPPD) and 170 a few miles away at Knowledge for Change (K4C). All feet were stored in cupboards at room temperature. The exact length of storage is unknown, but NCRPPD feet were older (1+ years) and not inspected prior to shipment while K4C feet were newer (<1 year old) and inspected prior to shipment.

The outer surfaces of the feet stored at NCRPPD and K4C were visually inspected by PM, and data were recorded/feet photographed by MB. Feet were manually manipulated to identify any flaws that the inspector considered likely to cause the foot to fail immediately or soon after being provided to the user (usability test). Occasionally, keels were removed from foot shells to identify prosthetic brand, but this was avoided when possible as keel removal can damage foot shells. Inner surfaces of foot shells were not inspected for this reason. Foot side (left/right), size, model (i.e., solid ankle cushioned heel – SACH – vs other), brand, and defects were recorded.

SACH feet derive their mechanical stiffness from both the harder inner keel and softer flexible surrounding material and can be used in modular prosthetics with the correct ankle attachment. “Other” feet were modular, often flexible keel prostheses, but included some axial, dynamic response (Energy Storing And Return; ESAR), and defunct hydraulic prosthetic feet. This category consisted of keels and generally removable foot shells, although some keels were integrated into foot shells in a way that prevented their removal. Brand model (e.g., Steeper’s Kinterra 3.0) was not recorded. No mechanical tests were run to investigate structural safety.

Multivariable binomial logistic regressions were run using the glm function in R/RStudio [37,38] to predict whether feet were usable (i.e., could be provided to prosthetic users). An initial regression was run including site of prosthetic storage, whether the foot was a left/right, foot size, foot model, and brand. A series of reduced statistical models were run testing each factor, independently, and a combination of independent variables. Models were compared using Akaike information criterion (AIC), and the best fit model was used for data interpretation.

## RESULTS

There were 196 left and 170 right feet ranging in size from 15 to 30 cm. 59 feet were SACH, and brand could be identified for 307/366 feet. The 307 feet represented 13 brands, the majority (60.66%) being Blatchfords (n = 120) or Ottobock (n = 102; Table 1; raw data in Electronic Supplementary Material, ESM). No damage was observed on any keels, but keels were not mechanically tested.

**Table 1:** Brands of feet available for prosthetic users in Fort Portal, Uganda. Sample (number usable, percent usable)

|  | BioQuest | Blatchfords | College Park Industries (CPI) | Fillauer | Janton | Kingsley Casual | Ossur |
| --- | --- | --- | --- | --- | --- | --- | --- |
| NCRPPD | 0 (0, NA%) | 58 (56, 96.55%) | 16 (12, 75%) | 4 (4, 100%) | 1 (1, 100%) | 2 (2, 100%) | 14 (9, 64.29%) |
| K4C | 1 (1, 100%) | 62 (62, 100%) | 11 (11, 100%) | 5 (5, 100%) | 0 (0, NA%) | 4 (4, 100%) | 11 (9, 81.82%) |
| Total sample | 1 (1, 100%) | 120 (118, 98.33%) | 27 (23, 85.19%) | 9 (9, 100%) | 1 (1, 100%) | 6 (6, 100%) | 25 (18, 72%) |
|  | Ottobock | Steeper | Streifeneder | Trulife | WillowWood | Unknown |  |
| NCRPPD | 59 (45, 76.27%) | 4 (4, 100%) | 2 (1, 50%) | 12 (8, 66.67%) | 7 (6, 85.71%) | 17 (15, 88.24%) |  |
| K4C | 45 (43, 95.56%) | 1 (1, 100%) | 0 (0, NA%) | 10 (8, 80%) | 1 (0, 0%) | 19 (16, 84.21%) |  |
| Total sample | 104 (88, 84.62%) | 5 (5, 100%) | 2 (1, 50%) | 22 (16, 72.73%) | 8 (6, 75%) | 36 (31, 86.11%) |  |

Binomial logistic regressions revealed site (NCRPPD vs. K4C) and brand were both important factors, and the statistical model that included these parameters. There is strong evidence that models including foot side, size, and model did not affect foot usability (dAIC > 4; Table 2). Statistical model m7 (weight = 0.915; Table 2), which only included parameters site and brand, was used for further analysis.

**Table 2:** Statistical models fit to the gathered data. **+** indicates the parameter was included in the model, while --- indicates the parameter was excluded. AIC values, difference in AIC (dAIC) values, and Weights were used to select the best fit model.

| **Model** | **Site** | **Side** | **Size** | **Model** | **Brand** | **AIC** | **dAIC** | **Weights** |
| --- | --- | --- | --- | --- | --- | --- | --- | --- |
| m7 | + | --- | --- | --- | + | 247.71 | 0 | 0.915 |
| m1 | + | + | + | + | + | 253.582 | 5.872 | 0.049 |
| m5 | --- | --- | --- | --- | + | 254.506 | 6.796 | 0.031 |
| m2 | + | --- | --- | --- | --- | 257.753 | 10.043 | 0.006 |
| m6 | --- | --- | --- | + | --- | 268.686 | 20.976 | 0 |
| m4 | --- | --- | + | --- | --- | 268.802 | 21.092 | 0 |
| m3 | --- | + | --- | --- | --- | 268.789 | 21.079 | 0 |

The feet stored at K4C were more usable than those stored at NCRPPD (Table 3). When accounting for the effect of brand (model m7), it was 102.5% more likely a foot stored at K4C was usable compared to NCRPPD (odds ratio = 2.025). The effects of brand on foot usability were examined to determine if certain brands might be more appropriate for donation than others, but as P&O centres may preferentially use certain brands, and P&O centre from which donations originated was not recorded, it is not possible to differentiate between these effects. Some brands were represented by few feet (i.e., <10) and the results for these should be ignored.

**Table 3:** Usable and broken feet per site.

| **Place** |  | **Count** | **Percent** |
| --- | --- | --- | --- |
| The Ninsiima Centre for the Rehabilitation of People with Physical Disabilities | Broken | 33 | 16.84 |
|  | Usable | 163 | 83.16 |
| Knowledge for Change | Broken | 10 | 5.88 |
|  | Usable | 160 | 94.12 |

When accounting for the effects of site and only considering brands with 10+ feet in the sample, Blatchfords feet performed the best, with 98.33% of the 120 feet being usable, overall (Table 4). This was followed by College Park Industries (CPI) and Ottobock with 85.19% and 84.62% usable feet, respectively. Trulife and Ossur performed the worst, with only ∼72% of their feet being usable, but the samples were small (<30 feet). The lack of effect of side, size, and model implies left/right and feet of any size were appropriate for donation. Similarly, SACH and “other” models were similarly appropriate for donation.

**Table 4:** Effect of brand on prosthetic foot usability. Coefficients taken from model m7 and measured relative to brand “BioQuest”. Higher values indicate a larger proportion of the feet were usable.

|  | Coefficient | Sample (percent usable) |
| --- | --- | --- |
| Janton | 1.107 | 1 (100%) |
| Steeper | 0.94 | 5 (100%) |
| Fillauer | 0.596 | 9 (100%) |
| Kingsley Casual | 0.471 | 6 (100%) |
| BioQuest | 0 | 1 (100%) |
| Blatchfords | -12.811 | 120 (98.33%) |
| College Part (CPI) | -15.061 | 27 (85.19%) |
| Unknown | -15.109 | 36 (86.11%) |
| Ottobock | -15.132 | 104 (84.62%) |
| WillowWood | -15.465 | 8 (75%) |
| Trulife | -15.911 | 22 (72.73%) |
| Ossur | -15.934 | 25 (72%) |
| Streifeneder | -16.459 | 2 (50%) |

Causes of foot failure included the foot shell having cracks, and being crumbly, discoloured, worn, or sticky due to polymer degradation (see Figure 1), with cracks being the most common cause of failure (Figure 2). Additionally, the keel could have been loose, or the foot could have been exceedingly stretchy. Of the 43 broken/unusable feet, three had two mechanisms of failure and the remaining 40 had one. Samples were not large enough to investigate the interaction between type of failure and prosthetic brand.

**Figure 1:**
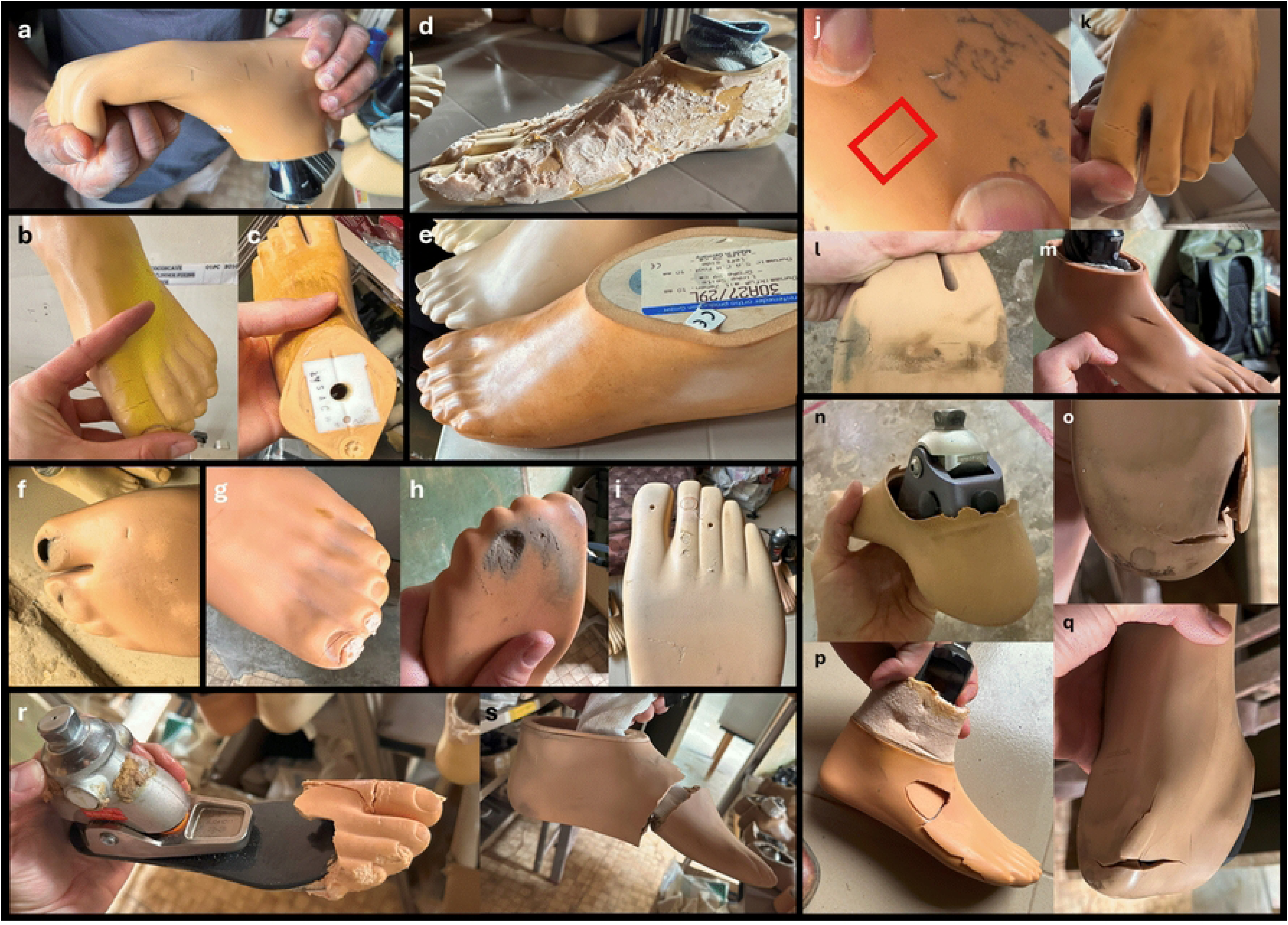
Examples of how feet could become unusable. a) polymer degradation, extreme flexibility in toes, b) discolouration and cracks on first toe, c) discolouration, d) a structurally stable (and therefore, usable) foot with cosmetic faults due to being covered in foam, e) degraded surface which became sticky to touch, f) foot shell worn through under toes, g-i) foot shells crumbling, j-m) small cracks, n-q) large cracks/fractures, r-s) catastrophic cracks/fractures causing mechanical failure.

**Figure 2:**
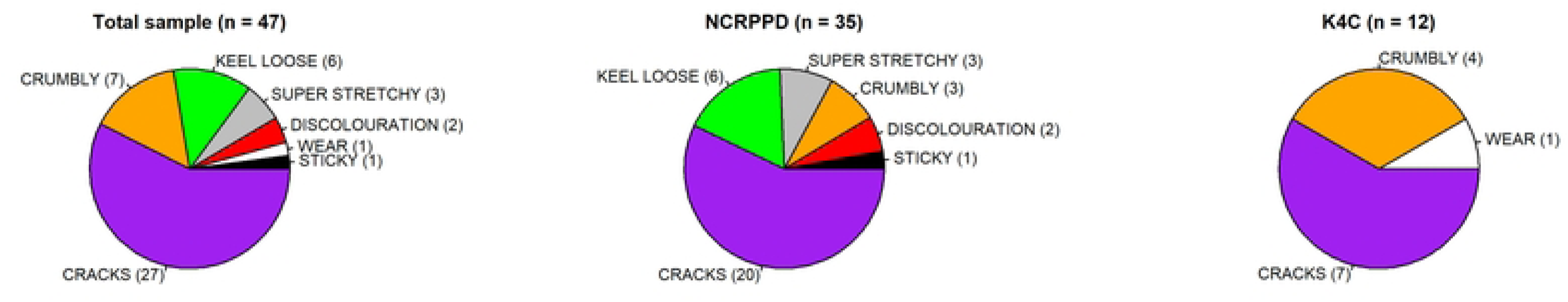
Issues which made prosthetic feet unusable, total and divided by centre.

When causes of foot failure were divided by centre (Figure 2), the feet at NCRPPD experienced all forms of failure except wear while the feet at K4C, which had gone through a usability inspection in Bristol, only failed due to cracks, crumbling, or wear. PM, who carried out the inspection for STAND in Bristol, stated the one foot with wear should have been removed using the inspection system in place, and its shipment to Uganda was user error.

Finally, differences in foot failure were examined between SACH and other feet, as SACH feet are commonly manufactured and provisioned in LMICs (e.g., by the International Committee of the Red Cross or Exceed Worldwide). SACH feet tended to become cracked, crumbly, or sticky, while other feet experienced all forms of foot failure.

## Discussion

Donated assistive technologies can be a major benefit to LMICs provided they meet standards and regulatory requirements. No (inter)national standards or regulatory requirements exist for used prosthetic devices, particularly those that are out of warranty. This can lead to substandard devices being supplied to LMICs, which can be a burden to the recipient LMIC [31,39,40].

> “[The] ‘Dumping’ of obsolete equipment by high-income countries (HICs) has been described as ‘morally reprehensible’ and has received adverse media attention. An ‘anything is better than nothing’ attitude, coupled with a donor–recipient power imbalance, has been cited the central reason for poor-quality donations. Recipients may be too embarrassed to point out the futility of donor efforts or may find it culturally inappropriate to decline a gift. As a result, ‘medical equipment graveyards’ of obsolete or broken donated biomedical equipment are commonly seen in hospitals across low-income and middle-income countries (LMICs) [31].”

To prevent morally reprehensible actions and the creation of prosthetic landfills, standards, like those in WHO’s principles and guidelines for the donation of medical equipment, can be applied to prosthetic devices [27,28,30]. The guidelines for donation clearly set out donor and recipient responsibilities for the donation of medical devices (Figure 2 in section 3.3; World Health Organization, 2024). The responsibilities of, e.g., understanding national regulatory requirements, conducting functionality tests, and submitting proof of functionality, lie with the donor, whereas the responsibility to refuse incomplete medical devices lies with the recipient. The Medicines & Healthcare products Regulatory Agency (MHRA) in the UK considers external prosthetic devices to be constructed of 2 parts, the socket and the “hardware” (i.e., all components up to the socket). Should recipients be refusing donations of prosthetic parts if they do not have access to the material (e.g., polypropylene) to manufacture sockets? Should donations of used prosthetic hardware be accompanied by the donations of raw materials to manufacture sockets? If so, how will this affect local markets for socket material, if present?

Ethical considerations extend past WHO’s principles, and must consider equitable, dignified, and inclusive prosthetic provision where, like in the WHO’s principles, the responsibilities of the donors and recipient are clearly stated. The equitable provision of prosthetic devices must be considered during device collection, checks, shipment, and dissemination (to the P&O centres and prosthetic user). At the point of collection, questions such as could the device be better used by the community it is being removed from? And could better or affordable versions of the same technologies be supplied new or manufactured locally in the recipient country?

All devices should be inspected, checked, and shipped to the same standard, regardless of donor or recipient. The WHO guidelines state that the donor and recipient should discuss quality and quantity and that the donated equipment should be functional for 2+ years post-donation for sustainable donation; however, a preponderance of lack of compliance with these guidelines has been found [41]. Strict compliance with the official WHO guidelines has been recommended for sustainable donation programmes involving medical devices and equipment [24,39,41,42].

Choice of country to ship devices to is important, not only considering the prosthetic user needs, but also the capabilities of the donors in those settings. Having an established network in a country with relatively lower need may trump providing devices to a country with a higher need but a worse network, as the former may ensure devices are supplied to users that need devices but the latter may not. Devices should be supplied to P&O centres in an equitable manner that reflects not only user needs, but the operation of the centre itself and to the patients on a need-basis, not first-come first-served [5].

The dignity of the recipients is paramount. As discussed previously, recipients may feel they must accept all donations in a grateful manner to ensure continued support [31]. This can be undignified for the recipients, who may feel the donations are not useful. Similarly, the devices themselves may have social or cultural issues (e.g., related to cosmesis) and prosthetic users may feel it is undignified to use these prosthetic devices, potentially leading to abandonment [10,12,43]. Although abandonment rates in higher income settings for some types of prosthesis and other assistive technologies are well-established [44–46], this issue has received little attention in other settings. Longer term, if donor organisation were required by their funders/supporters to report not only the number of prostheses donated, but also the abandonment rates, this might focus attention on appropriateness of donations as well as the quantity. Social, cultural, etc., factors [4] must be considered when ensuring dignified provision. As these devices are basic human rights, users should not be made to feel they are fortunate or lucky to receive the devices.

Obligations of the donors should be considered across time scales. On the short scale, donors should at a minimum ensure they are providing technologies that work, can be used by the recipient (correct shape, size, and provisioned by prosthetists/technicians who are trained in that technology), are fulfilling a need, and are wanted by the recipient. On the long scale, donors should ensure the systems are robust and work well [13,29,31]. When creating a new supply chain (i.e., donations), it is the ethical responsibility of the donors to understand what supply chains they are replacing and how they will ensure supply chains continue when the donations stop, ensuring the system is sustainable and does not prevent local innovation [5,6]. Given the manufacturer of devices will likely only provide user manuals in languages relevant to their key export markets, careful consideration should be given to how such information can be made accessible to recipients in other countries [34].

Clear standards exist for the design prosthetic design (e.g., ISO 10328 and 22675), manufacturing (e.g., ISO 9001), and dissemination (e.g., WHO prosthetics should be available for all). “Poor quality assistive products exist due to inadequate standards, lack of regulatory enforcement and lack of knowledge about the need for safe and effective products [20].” What standards/regulatory requirements should exist for prosthetic donation/provision?

If donated prosthetics are new and not yet used, they should meet the standards for prosthetic provision of the donor country. But if used, separate standards need to be applied.

### A proposed set of standards for the donation of used prosthetic feet

Here, we propose and evaluate a set of standards to govern the donation of prosthetic feet. These are consistent with many of the WHO’s indicators of suitability for medical device donation (i.e., appropriate for setting, assured quality and safety, affordable and cost-effective; [27]). Also consistent with many attributes of the WHO’s principles of good donation, we suggest it is the responsibility of the donor to carry out these checks (i.e., conducting functionality tests) before the prosthetic feet are donated to the recipient, and that some proof of functionality (i.e., submitting proof of functionality) should be submitted to the centre for each prosthetic foot [27,28,30].

Currently, we suggest these standards be taken on as internal operating procedures by donors, and stress that the adherence to these standards is the responsibility of the donors and not the recipients. In the future, we recommend any future alterations, additions, or substitutions to these standards should also incorporate structural testing and potentially simplified versions of quality assurance tests carried out by manufacturers. We further recommend the creation of formal, international regulatory requirements/standards to govern prosthetic donation be created, to ensure quality and prosthetic safety.

STAND’s internal inspection protocol increased the quality of prosthetic foot provision and led to a 65.1% reduction in unusable feet donated to Uganda. However, unusable prosthetic feet that were cracking/crumbling were still being donated (Figure 2), possibly because feet were missed during inspection or were experiencing further degradation during storage and/or travel. The inclusion of a chemical or mechanical polymer testing for prosthetic foot shells may be beneficial. An altered form of the inspection method, informed by results here, is presented in Table 5.

**Table 5:** Suggested checklist to ensure quality and standards for donation of prosthetic feet.

| Checklist questions | Logic/reasoning |
| --- | --- |
| 1) Is the foot complete? (Fig 1r) | Incomplete feet are not usable. While parts can be used to fix broken feet, it is better practice to supply complete feet rather than parts for feet the P&O centre may not have |
| 2) Are there any flaws related to mechanical function?<br>Examples include stripped screws, broken keels, cracks of any size and surface degradation (e.g., stickiness, crumbling; Fig 1e, Fig 1f-s) | Mechanical failures will prevent the prosthetic from being usable. Cracks or surface degradation, like stickiness or crumbling, can indicate polymer degradation and that the foot has mechanically failed or will begin to mechanically fail soon. This is particularly true for feet with removable foot shells. |
| 3) Is the foot compliant? (Fig 1a) | If too pliable, the foot will not be rigid enough to support a person's weight during locomotion and is prone to break |
| 4) Are there any surface for flaws related to cosmesis?<br>Examples include surface degradation (e.g., crazing; Fig 1c), discolouration (Fig 1b), inclusion of other materials (Fig 1d), and staining | Cosmetic issues can lead to prosthetic abandonment |
| For feet with removable foot shells |  |
| 5) If the foot has bumpers, are they present? | Bumpers change foot stiffness, improve alignment, and enable smooth gait |
| 6) Are keels and foot shells correctly matched? | Incorrect matches between keels and foot shells can cause poor fits, leading the shell to move relative to the keel and/or detach during use |

On the checklist, factors related to mechanical function should be treated as hard cut-offs; prosthetics that cannot guarantee the user’s physical health and/or will break soon after donation should not be provisioned. Factors related to cosmesis (e.g., Fig 1d) require judgement from the provisioner, and recipients should be allowed to turn down donations for cosmetic reasons [31] as they could cause psychological, social, and/or cultural issues. Of note, cosmetic factors, some of which are related to prosthetic acceptance and abandonment, such as prosthetic colour, are largely missing from Table 5. Similarly, economic, social, cultural, etc., factors [4] are not considered here, but should be included in prosthetic donation standards/regulations. Investigations into quality assurance checks by prosthetic manufacturers may help improve quality standards for used prosthetics. The co-development of checklists with both donors and recipients may help improve donations [39].

### Repair, reuse, recycle, and systems surrounding the donation of used prosthetic feet

Prosthetics can last long in LMICs [47–49]. Prosthetics in HICs are overdesigned for their intended length of use, making them prime for repair, reuse, and recycle. In the UK, prosthetics are often replaced due warranties or guidelines and not prosthetic breakages (personal communication). Implementing a repair and reuse model is critical for prosthetic donation and can help promote prosthetic longevity in LMICs, particularly in rural areas, which have relatively more amputees, lack P&O centres, and are difficult to reach [2,3,8,18]. Different barriers exist in implementing a repair, reuse, recycle model for prosthetic provision within and between LMICs. For example, in India, cost of was the biggest barrier for upper limb body-powered prosthetic repair [43]. In Uganda, the biggest barrier to repair differs depending on who is consulted; from the prosthetist’s perspective, it is materials, but from the patient’s perspective, it is cost [5].

In this way, an integration of global and local markets, where prosthetics are supplied internationally and repaired locally, may be an efficient way forward [6,20]. Global markets can drive down the cost of prosthetics due to bulk manufacturing – assuming prosthetics can be purchased at a level to drive down the cost [20] – but are likely to be disrupted by global crises, like the Covid-19 pandemic [6]. The combination of many different technologies, requiring large levels of stock for spare parts and large skillsets ranging all possible products, however, may be problematic for local prosthetic repair. Staff shortages, large patient numbers, lack of components, and practical and financial challenges mean prosthetists in LMICs need to be more creative in technology creation and provision and likely require more training and skillsets than prosthetists in HICs [4]. Modular prosthetics make the integration of different technologies easier, and reduces the necessary stock, but limits product variability, particularly with lower-limb prosthetic cosmesis. The ability to manually construct and repair prosthetics is important to successful prosthetic provision under the repair, reuse, and recycle model [2,6].

The integration of digital manufacturing and 3D printing into prosthetic provision and repair could aid in the local manufacture of prosthetic parts, reducing the volume of spare parts that must be kept in stock and allowing the manufacture of parts for the repair of older devices that are no longer being manufactured. The digital manufacture of parts requires the establishment of local manufacturing centres, upskilling workers, creation of supply chains for both manufacture of prosthetics and maintenance of equipment, 3D digital models of the parts to be manufactured – which organisations may not be willing to share – as well as the construction of adequate manufacturing facilities (i.e., rooms with consistent, reliable power operating within tolerable temperatures).

A systematic review on the application of 3D printing to prosthetics in developing countries was conducted in 2021 [50]. They identified 14 studies from 2014-2020 that included a total of 47 prosthetic users, where devices were not always manufactured or tested in the developing country. The biggest advantages to 3D printing prosthetics were the ability to manufacture low-cost prostheses without loss of functionality and the ability to fit prosthetics at a site other than the place of manufacture. Prosthetics were generally cheap, but not always so. The biggest drawbacks were time to manufacture and that most studies did not conduct trials in LMICs. Not discussed are issues related to local infrastructure [31], supply chains and related trade barriers [51], maintenance and repair of equipment, etc. “…unreliable supply of quality prosthetic devices and components is a significant hindrance, slowing or stopping fitting [11]”. It was also unclear in the papers reviewed what parts were 3D printed, what the lifespan of the devices were, etc., [50]. It appears that, as far as 3D printing prosthetics in LMICs is concerned, device design is a focus, and most other related factors are ignored [52].

Reusing prosthetics can lead to a reduction of waste, but inappropriateness or user dissatisfaction in assistive technologies can lead to abandonment, increasing waste [6]. In Norway, 1/3 of assistive devices are reused through the Norwegian Assistive Technology Provision Model [6]. In other HICs re-use rates are highly likely to be lower; for example, the UK NHS has recognised that many walking aids do not get re-used [53]. However, technologies are not always appropriate to reuse in LMICs, and the “something is better than nothing” paradigm is not always true [54]. Time should be given to study and understand the local environment and culture [4] – devices should be low cost, locally available, capable of manual fabrication, considerate of local climate and working, durable, simple to repair, simple to process using local production, reproducible by local personnel, technically functional and not too high-tech, biomechanically appropriate, lightweight, and cosmetic, psychosocially appropriate [4].

### Future work

The use of donated, used prosthetic limbs and their components has the potential to be a major source of prosthetic supply. Donors should adopt minimum standards to ensure device quality and function, devices fulfil end-user needs (e.g., biomechanical, social, cultural), and devices work within local systems without displacing existing services. International standards to ensure quality should be created, potentially with an associated accreditation process operating within a regulatory environment.

To begin this journey, we proposed a set of standards in the form of a quality assurance check that focus on prosthetic foot quality at the point of provision (Table 5). However, these standards do not test product safety or lifespan. Rapid, affordable, and non-/minimally destructive tests should be created to test prosthetic foot safety/lifespan to promote prosthetic user safety and ensure users will not need to have their devices repaired/replaced soon after receipt. Some test(s) should focus on the structural integrity of the keel, potentially similarly to the ISO 10328 and 22675 tests. Given the issues observed with the foot shells (Figure 2), non-/minimally destructive methods for quantifying polymer quality should be developed/applied to prosthetic feet to further provide adequate prosthetics to prosthetic users in LMICs. Critical data are needed to shape these standards/tests, including:

- When/why are feet being replaced at the time of collection for reuse and recycle?
- How long do feet last once repaired and provisioned?
- What factors affect used prosthetic foot safety and lifespan?
- What differences exist in user quality of life between new and used prosthetic feet?

Finally, it is possible different standards will be required for different foot models because of differences in failure mechanisms. For example, SACH feet, where the keel is fully integrated into the prosthetic foot, did not suffer from keel loosening, but other prosthetic foot models did (Figure 3).

**Figure 3:**
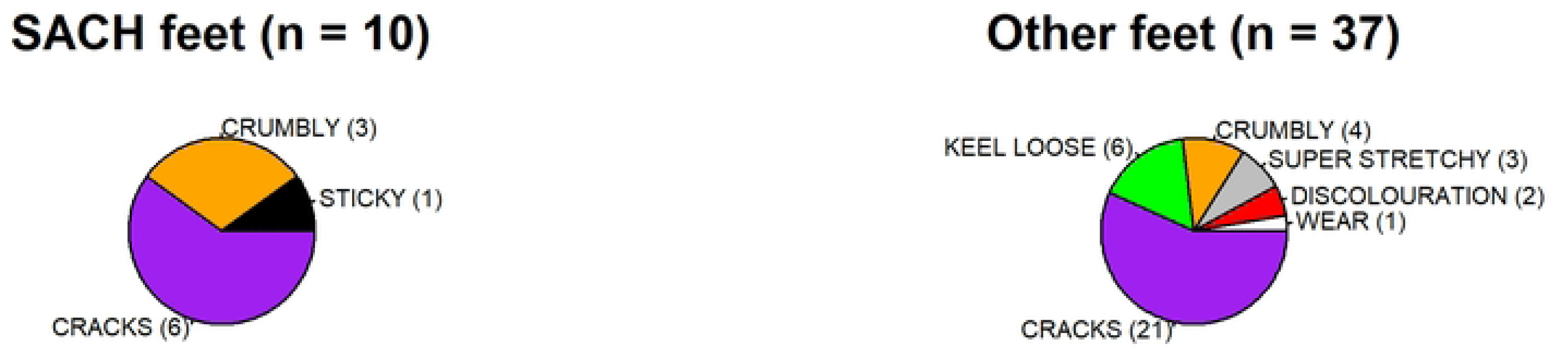
Issues with SACH vs other prosthetic feet.

### Limitations

A lack of consistency in brands, and knowledge of models or where the prosthetic feet originated from, means it is not possible to make conclusions about which brands are better/worse for donation. While ∼400 feet is a large sample, it is also not large enough to determine e.g., the effect of brand on usability, as the sample sizes for many brands are too small.

The feet at the NCRPPD have been stored at ambient temperature in Fort Portal for longer than those at K4C. Polymers, like the foot shells, degrade under warmer temperatures, which is why it is recommended prosthetic feet be stored at certain temperatures. However, the range of storage temperatures for prosthetic feet can be vast (e.g., −15 to 50°C for Blatchfords ELAN feet, models ELAN22L1S—ELAN30R8S and ELAN22L1SD—ELAN30R8SD) and, while temperature was not recorded in the storage facilities at these centres, it is unlikely to have exceeded 50°C. It is therefore possible some of the differences we observed in foot failure were due to differences in storage time in warm temperatures.

## Conclusions

Here, we propose a set of standards for the donation of used prosthetic feet. We have demonstrated the efficacy of these standards, and hope is these standards will lead to improved and consistent quality in donated prosthetic feet, particularly for LMICs. We stress that the responsibility of adhering to these standards lies with the donor and not the recipient. These standards do not consider many aspects important for prosthetic feet (e.g., structural stability) which should be considered in future additions/alterations to this methodology. The creation of standards/regulatory requirements governing the donation and provision of used prosthetics is critical not only for the donation of prosthetics to LMICs, but the creation of a circular economy surrounding prosthetic provision in HICs.

## Data Availability

All data is available as ESM

## Acknowledgements

We would like to thank STAND, Knowledge for Change, The Ninsiima Centre for the Rehabilitation of People with Physical Disabilities, British Academy for working with us on this project and/or funding this research. We would like to thank members of STAND for information on the carbon footprint of their work and for comments on data collection/analysis. We would further like to thank Prof Nachiappan Chockalingam for comments on an early version of this manuscript.

## Notes

### Competing Interest Statement

One author, Ackers, runs K4C, and one author, Promise, works for STAND

### Clinical Trial

NA

### Funding Statement

This research was partially funded by Knowledge for Change (K4C)

